# Baseline predictors of mortality in non-idiopathic pulmonary fibrosis interstitial lung disease – A retrospective cohort study at a tertiary centre in Malaysia

**DOI:** 10.64898/2026.02.12.26346139

**Authors:** Leng Cheng Sia, Chee Kuan Wong, Dinesh Sivakumar, Deevan Marc Chandran, Kee Ying Yeoh, Shan-Yan Ling, Fui Bee Woo, Guat Yee Lim, Wai Ling Leong, Mau Ern Poh, Yong-Kek Pang

## Abstract

**Background:** Baseline factors associated with mortality in non-idiopathic pulmonary fibrosis interstitial lung disease (non-IPF ILD) have been described in several population, but data from Malaysian cohorts remain limited.

**Objectives:** We described the clinical characteristics and explored baseline factors associated with all-cause mortality in a single tertiary centre in Malaysia.

**Methods:** This retrospective cohort study analysed non-IPF ILD treated at a tertiary centre in Malaysia between 1 January 2010 and 31 May 2023. Patients without baseline lung function tests or high-resolution computed tomography were excluded. Survival was assessed using Kaplan– Meier analysis, and associations with mortality were explored using Cox regression.

**Results:** The cohort included 229 patients, with a mean age of 60 ± 15 years, and a male-to-female ratio of 1:3. Indian patients constituted the largest group (n = 109, 47.6%). The mean baseline forced vital capacity (FVC) was 53.3 ± 21% predicted. Older age, FVC <50% predicted and idiopathic interstitial pneumonia were associated with higher mortality in reported multivariate analysis. Compared with Malay patients, hazard ratios were 9.86 (95% CI 1.27– 76.89) for Chinese patients and 8.59 (95% CI 1.14–64.69) for Indian patients, although the overall adjusted test for ethnicity was not statistically significant (p = 0.091). Survival differed significantly across non-IPF ILD subtypes (log-rank p = 0.048). Patients with hypersensitivity pneumonitis had the shortest estimated median survival in this cohort (6.4 years).

**Conclusion:** Ethnicity showed a potential association with mortality in this single-centre non-IPF ILD cohort. Larger prospective studies are needed to confirm this association.

## Introduction

Interstitial lung disease (ILD) comprises a heterogeneous group of disorders, characterised by inflammatory and/or fibrotic changes in the lung parenchyma. (1) Non-IPF ILDs include a wide spectrum of aetiologies such as connective tissue diseases, drugs or environmental agent exposure, pulmonary sarcoidosis, and unknown causes. Previous studies have identified age, lung function, extent of fibrosis, and ILD subtype as factors associated with survival. The gender–age–physiology (GAP) model has been validated for predicting IPF mortality based on baseline characteristics, and adaptations including ILD-GAP have been evaluated for non-IPF ILD, such as rheumatoid arthritis RA-ILD, systemic sclerosis SSc-ILD, unclassifiable ILD, and mixed cohort. (2, 3) However, the applicability of reported associations to individual referral populations requires careful assessment.

Data on baseline mortality predictors in non-IPF ILD from our region are limited. Therefore, we aimed describe patients with non-IPF ILD attending a Malaysian tertiary centre and to explore associations between baseline characteristics and all-cause mortality.

## Methods

### Study design and study population

This retrospective cohort study included patients with non-IPF ILD treated at the Universiti Malaya Medical Centre (UMMC) from 1 January 2010 to the censor date of 31 May 2023. UMMC is a tertiary and referral centre for ILD, located in Kuala Lumpur, the capital of Malaysia. It covers a 243 km^2^ area and has a population of 1.77 million, with a population density of 7299 per sq. km. Malays and Chinese make up most of the population (46.5% and 42.4% respectively), followed by Indians (9.8%) and others (1.3%). (4)

### Ethics

This study was approved by the hospital’s Medical Research Ethics Committee on 12 October 2024 (MECID. No 20241012-14306) and informed consent was waived.

### Inclusion and exclusion criteria

Adult patients (≥ 18 years) with a diagnosis of ILD were identified from the hospital registry using the relevant ILD codes listed in the International Classification of Diseases, Tenth Revision (ICD-10) codes, including J84 and D86. Patients without baseline HRCT or RFT data were excluded because imaging and lung physiology were required for study assessment. Pulmonary fibrosis unrelated to ILD and non-pulmonary sarcoidosis were excluded. The cohort also included rare lung diseases such as lymphangioleiomyomatosis, pulmonary aluminosis, pulmonary amyloidosis and diffuse cystic lung disease. All ILD cases were cross-checked by experienced pulmonologists (YKP, LCS) and a thoracic radiologist (WYL), and cases not fulfilling the International Guideline of Diagnosis of ILD were excluded. (5–7)

### Data collection

Clinical records were reviewed using the electronic medical record, and radiological images were reviewed using the picture archiving and communication system. Demographic characteristics, smoking history, occupation, medications, comorbidities, allergies and family history were captured in a standardised electronic spreadsheet. Baseline investigations such as RFT, lung biopsy, bronchoscopy and autoantibody results obtained at diagnosis, were also recorded.

### Imaging and lung function test

HRCT scans were obtained with <1.5 mm collimation at full inspiration and reconstructed using high-resolution algorithms. (8, 9) Lung function tests were performed in the clinic using portable spirometry or in the laboratory according to the European Respiratory Society recommendations (10)

### Outcome and follow up

The outcome was all-cause mortality. Survival time was calculated in years from the date of diagnosis to death. The date of diagnosis was defined as the date of HRCT confirming fibrosing ILD. Patients who remained alive and those lost to follow-up were censored on 31 May 2023.

### Data analysis and definition

Categorical variables were summarised as percentages, while continuous variables were summarised as the mean ± standard deviation (SD), median or range, depending on the normality of distribution of the variables. Categorical variables were compared by using the chi-squared test or Fisher’s Exact test, as appropriate. Continuous variables were compared using one-way ANOVA or the Kruskal-Wallis test.

Cox proportional hazards regression was used to explore associations between baseline characteristics and all-cause mortality. In the reported analysis, variables with univariable p < 0.25 were considered for inclusion in the multivariable model (11, 12) Associations were expressed as hazard ratios (HRs) with 95% confidence intervals (CIs). A two-sided p <0.05 was the nominal threshold for statistical significance. Analyses were conducted using SPSS version 26. The findings were interpreted as exploratory associations rather than as validated individual mortality predictions.

## Results

### Basic characteristic of ILD patients

A total of 229 patients were included (Figure 1). Baseline demographic and clinical characteristics are summarised in Table 1. Distribution of non-IPF ILD subtypes is shown in Figure 2. The female-to-male ratio was approximately 3:1, and CTD-ILD predominated. Indian patients constituted the largest ethnic group (n = 109, 47.6%).

**Fig 1.**
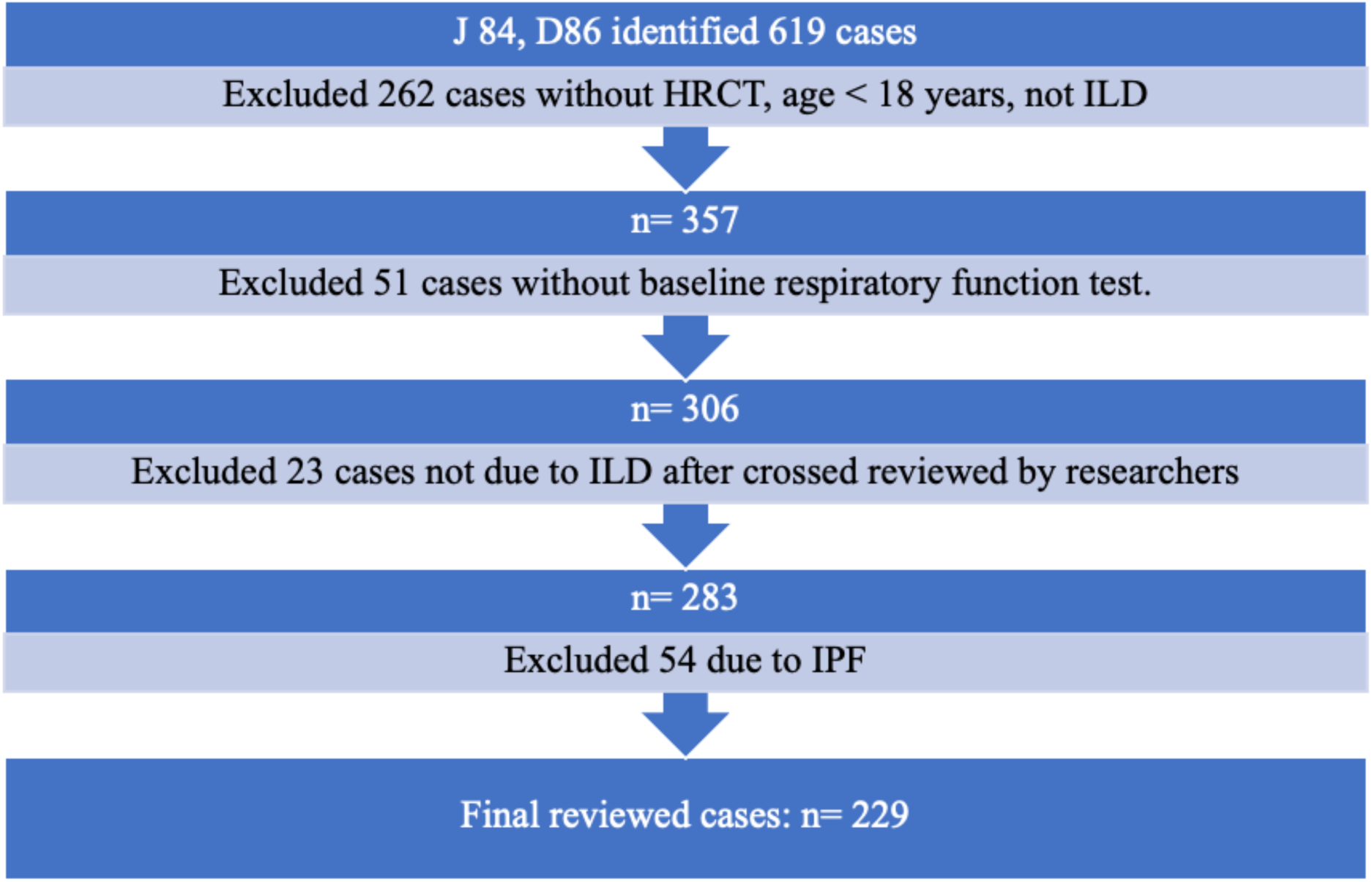
Flow chart of case selection. HRCT= High-resolution computed tomography; ILD= Interstitial lung disease

**Fig 2.**
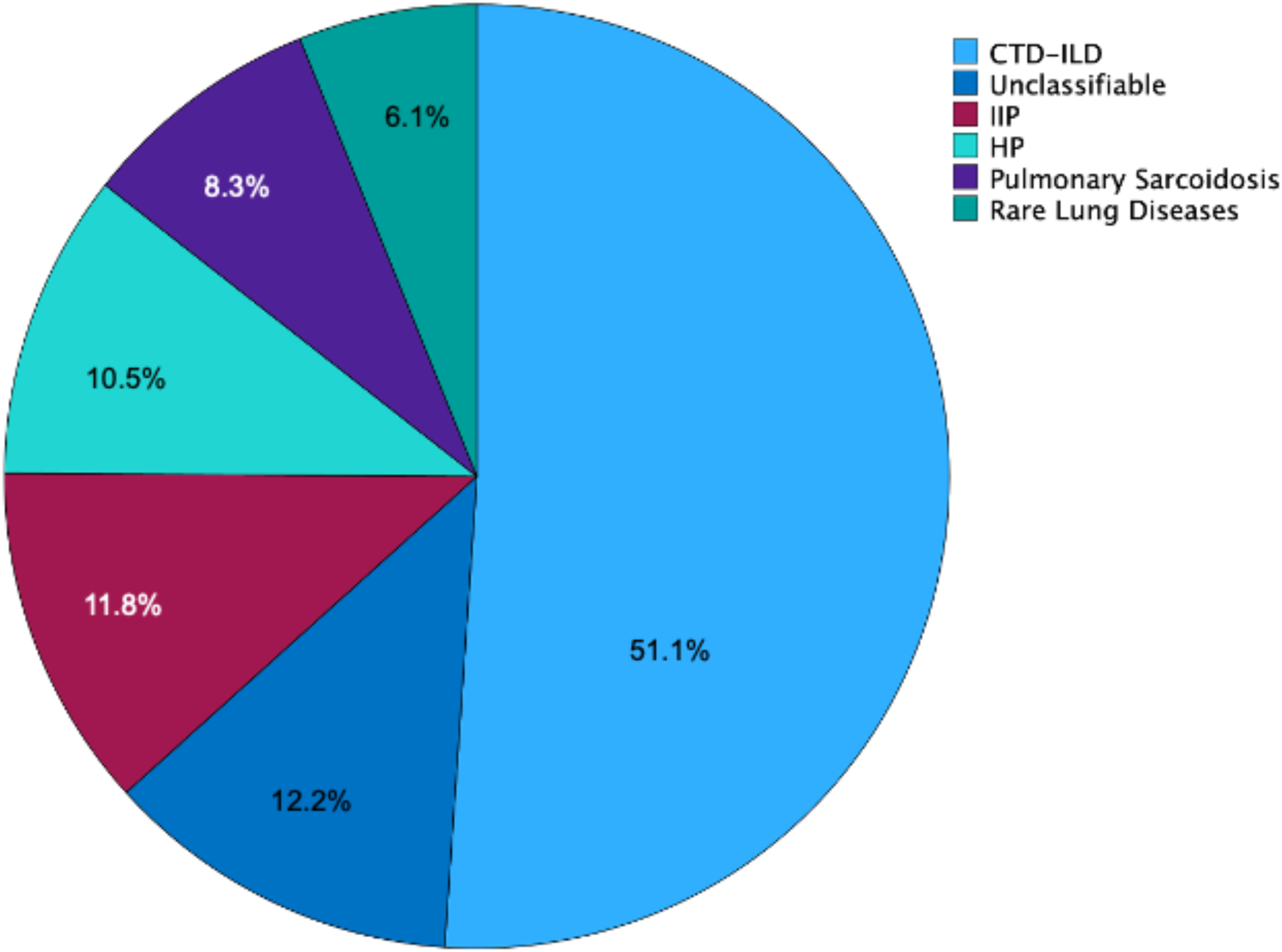
Proportion of ILD subgroups. CTD= connective tissue disease; IIP = idiopathic interstitial pneumonia; HP = hypersensitivity pneumonitis

**Table 1.**
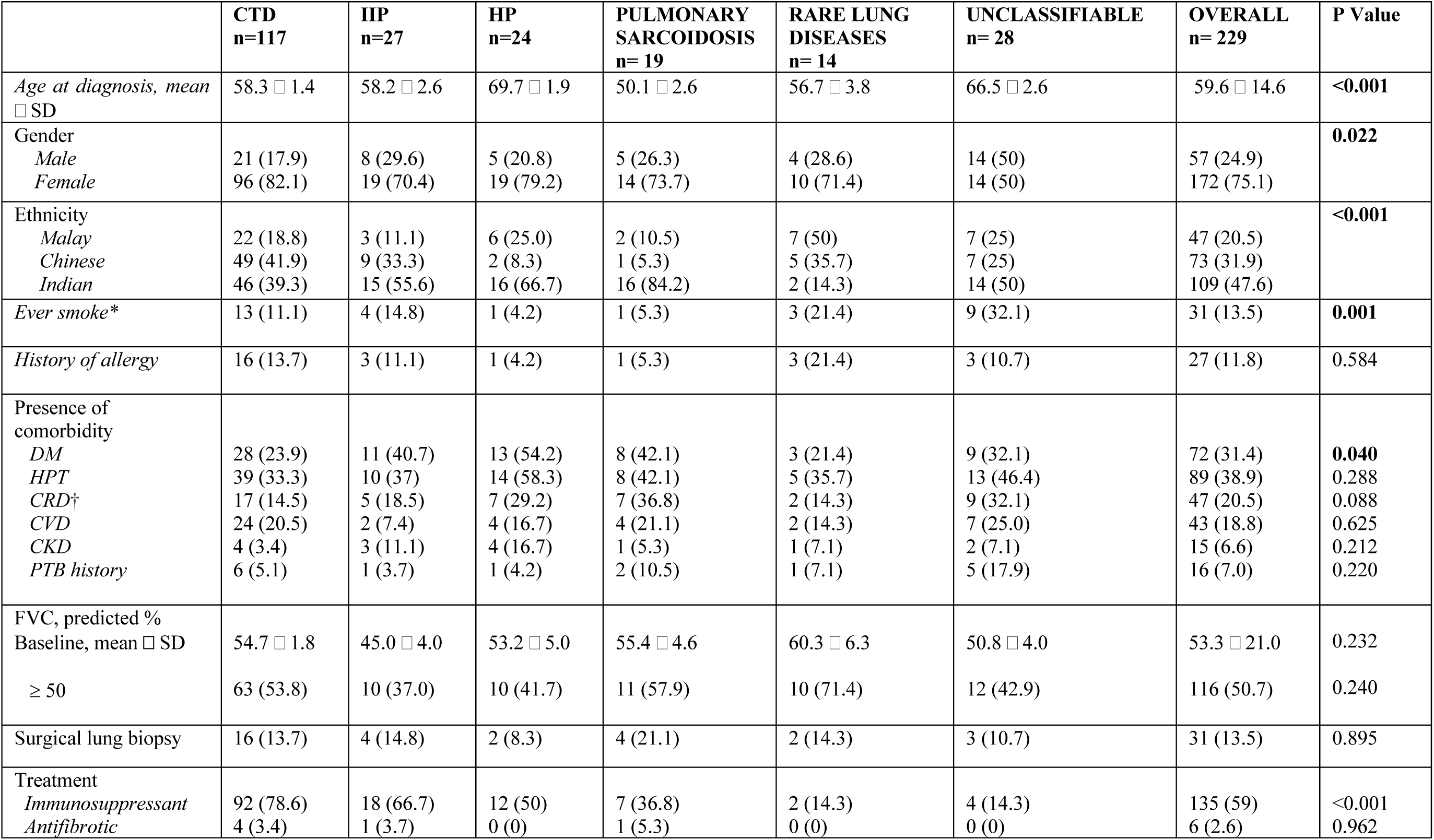
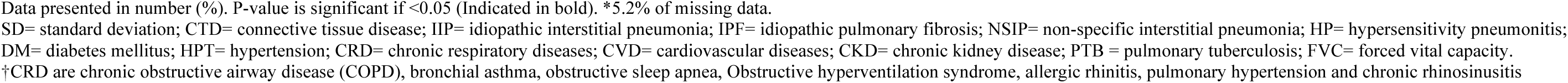
Sociodemographic, baseline clinical characteristics, investigations, and treatment of patients with non-IPF interstitial lung disease.

### Factors associated with overall mortality

Univariate and multivariate Cox regression findings are presented in **Table 2**. In multivariable model, age ≥ 50 years and FVC <50% were associated with higher mortality (HRs of 3.23, P = 0.032; and HR 3.65, p = 0.001, respectively). Compared with Malay patients, the adjusted HR was 9.86 (95% CI 1.27 – 76.89, P = 0.029) for Chinese patients and 8.59 (95% CI: 1.14–64.69, P = 0.037) for Indian patients. Compared to CTD-ILD, IIP was associated with higher mortality (HR 2.39, P = 0.048).

**Table 2.** Factors affecting mortality in non-IPF ILD (Cox regression analysis)

| Variable | Univariate analysis |  |  | Multivariate analysis |  |  |
| --- | --- | --- | --- | --- | --- | --- |
|  | HR | 95% CI | P-value | HR | 95% CI | P-value |
| Age≥ 50 | 3.64 | 1.28-10.37 | **0.016** | 3.23 | 1.11-9.43 | **0.032** |
| Female | 0.63 | 0.31-1.29 | 0.208 | 0.83 | 0.38-1.79 | 0.634 |
| Ethnicity |  |  | 0.074 |  |  | 0.091 |
| Malay | 1 | - | - | 1 | - | - |
| Chinese | 8.89 | 1.15-68.38 | **0.036** | 9.86 | 1.27-76.89 | **0.029** |
| Indian | 10.31 | 1.39-76.51 | **0.023** | 8.59 | 1.14-64.69 | **0.037** |
| Smoking history | 0.63 | 0.19-2.07 | 0.450 | - | - | - |
| FVC <50% of predicted | 3.31 | 1.59-6.91 | **0.001** | 3.65 | 1.65-8.06 | **0.001** |
| Comorbidities |  |  |  |  |  |  |
| DM | 1.20 | 0.60-2.43 | 0.603 | - | - | - |
| HPT | 1.32 | 0.67-2.61 | 0.424 | - | - | - |
| CRD | 1.19 | 0.54-2.61 | 0.671 | - | - | - |
| CVD | 1.29 | 0.59-2.85 | 0.525 | - | - | - |
| CKD | 0.97 | 0.23-4.06 | 0.973 | - | - | - |
| PTB history | 0.37 | 0.05-2.69 | 0.323 | - | - | - |
| ILD subtypes |  |  | 0.091 |  |  | 0.270 |
| CTD | 1 | - | - | 1 | - | - |
| IIP | 2.62 | 1.12- 6.13 | **0.027** | 2.39 | 1.01-5.68 | **0.048** |
| HP | 2.55 | 0.90- 7.17 | 0.077 | 2.10 | 0.70-6.28 | 0.185 |
| Pulmonary Sarcoidosis | 0.27 | 0.03- 2.02 | 0.200 | 0.36 | 0.05-2.85 | 0.335 |
| Rare lung diseases<sup>†</sup> | 0.78 | 0.10-5.90 | 0.811 | 1.41 | 0.17-11.67 | 0.751 |
| Unclassifiable | 1.48 | 0.49- 4.44 | 0.485 | 1.14 | 0.35-3.71 | 0.822 |

N= 229. P-value is significant if <0.05 (Indicated in **bold**).
HR= hazard ratio; CI= confidential interval; FVC= forced vital capacity; DM= diabetes mellitus; HPT= hypertension; CRD= chronic respiratory diseases; CVD= cardiovascular diseases; CKD= chronic kidney disease; CTD= connective tissue disease; IIP= idiopathic interstitial pneumonia; HP= hypersensitivity pneumonitis.
<sup>†</sup>lymphangioleiomyomatosis, pulmonary aluminosis, pulmonary amyloidosis and diffuse cystic lung disease.

### Survival by ILD subtype

Kaplan–Meier analysis in **Figure 3** showed differences in survival across non-IPF ILD subtypes (log-rank p = 0.048). HP had the lowest median survival (6.4 years); however, only two patients remained at risk when the median was reached, limiting the precision of this estimate. CTD-ILD and pulmonary sarcoidosis had more favourable observed survival.

**Figure 3.**
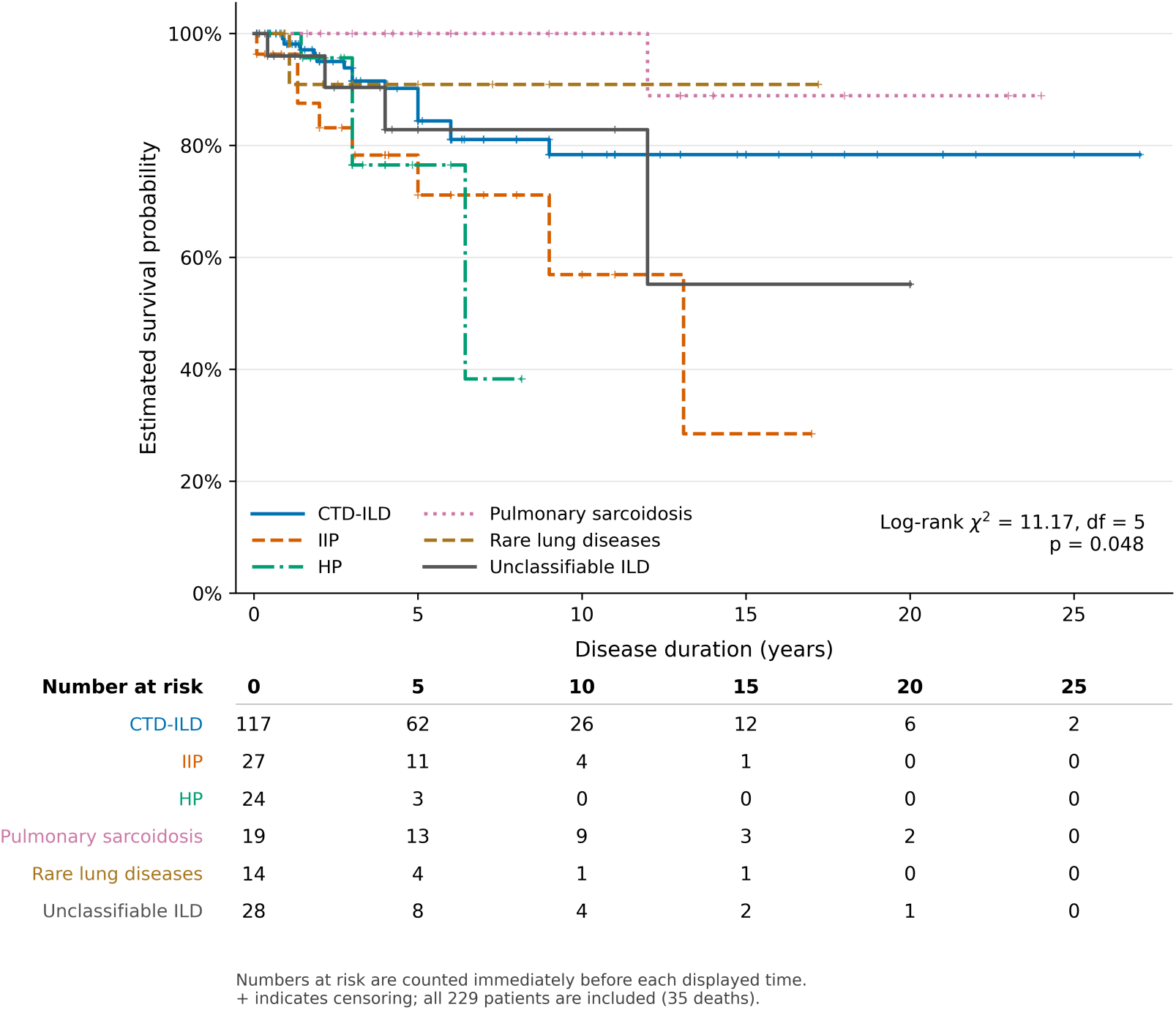
Kaplan-Meier survival curve for non-IPF ILDs. CTD= connective tissue disease; IIP= idiopathic interstitial pneumonia; HP= hypersensitivity pneumonitis

## Discussion

In this single-centre, multi-ethnic Malaysian cohort of non-IPF ILD, Indian patients disproportionately represented despite comprising a minority of the regional or national population. Age ≥50 years and baseline FVC <50% predicted were associated with higher adjusted mortality hazards. Chinese and Indian patients had higher adjusted mortality hazards than Malay patients in individual comparisons. Survival differed across ILD subtypes, with HP having the shortest estimated median survival, while pulmonary sarcoidosis and CTD-ILD showed favourable observed survival. These findings highlight the prognostic relevance of baseline physiology and ILD subtype, also suggest that ethnicity warrants further investigation.

Our findings are consistent with previous work demonstrating the importance of age, physiological impairment and ILD subtypes in prognostic assessment. Age and baseline lung physiology have been shown to affect mortality in non-IPF ILD. (13–15) The PROGRESS study identified age ≥50 years as a mortality-associated factor and reported poorer survival among patients with unclassifiable ILD, IIP or HP than among those with autoimmune ILD. (15) Ryerson et al. evaluated the GAP model across 1,012 patients with chronic ILD and developed the ILD-GAP index, incorporating disease subtype alongside gender, age and lung physiology. (2) These similarities support the continued clinical value of baseline assessment alongside longitudinal monitoring for progression.

A notable difference from other cohorts was the degree of baseline physiological impairment. Mean FVC in our cohort was 53.3 ± 21% predicted, compared with 62.1 ± 17.4% in the Singaporean non-IPF fibrotic ILD cohort reported by Kam et al. and 74 ± 22% in the French

PROGRESS cohort. (15, 16) These comparisons suggest that our patients had greater average physiological impairment at assessment. Differences in study populations, referral pathways, timing of presentation, ILD subtype distribution and lung function reference equations may contribute to the variation. In our cohort, FVC <50% predicted was associated with higher mortality, whereas an exploratory 60% cutoff produced a less conclusive adjusted estimate (S2 Table). This is consistent with the Japanese prospective registry reported by Tsubouchi et al., in which patients with non-IPF idiopathic interstitial pneumonias and FVC <50% predict poorer survival than those with higher FVC, and Imai et al. who demonstrates that baseline FVC <50% is associated with poorer transplant-free survival even among patients with physiologically stable non-IPF ILD.(17, 18) These findings support the prognostic relevance of baseline physiological impairment across different non-IPF ILD populations.

Chinese and Indian patients had higher adjusted mortality hazards than Malay patients in individual comparisons. However, the overall adjusted test for ethnicity did not reach statistical significance (p = 0.091), and the estimates had wide confidence intervals. These findings suggest possible ethnic differences in mortality but remain exploratory. Relevant regional evidence comes from the Singaporean cluster analysis by Kam et al., which identified a predominantly Chinese cluster with survival similar to IPF and a predominantly Indian cluster with the highest mortality and lowest baseline lung function. (16) Although that study assessed combinations of characteristics rather than an independent ethnicity effect, it provides complementary evidence that ethnic composition may accompany clinically meaningful differences in non-IPF ILD phenotypes. African Americans have been reported to experience higher mortality in sarcoidosis but favourable survival in IPF, providing an interesting comparison. (19, 20) In addition, CTD-ILD has been reported to have poorer outcomes among Black patients, especially for ILD associated with systemic lupus erythematosus and systemic sclerosis. (21, 22) Potential explanations include differences in exposure profiles, disease presentation, comorbidities, socioeconomic circumstances and access to specialist care. These remain hypotheses rather than mechanisms established by our analysis. Our findings therefore provide a rationale for investigating ethnicity together with these clinical and contextual factors in larger regional cohorts.

In survival analyses, HP had the shortest median survival of 6.4 years, approaching the reported median survival of 2-5 years in IPF. (23) This broadly aligns with the PROGRESS and the Canadian registry, supporting the potentially aggressive course of fibrotic HP. (13, 15) Separately, fibrotic bronchiolocentric interstitial pneumonia (previously known as HP) imaging pattern was associated with the poorest survival within CTD-ILD. (24) Conversely, a Hong Kong study reports a substantially longer median survival of 11 years of HP, similar to a German registry reporting a median survival of 16 years. (14, 25) These differences may reflect variation between cohorts in demographic and ethnic composition, baseline disease severity, and environmental exposures. For instance, HP is recognised as the most common ILD subtype in India. (26)

Several limitations should be recognised. First, the retrospective design may have introduced selection and information bias during case identification and data collection. Patients without baseline lung function data (n = 51) were excluded and may have differed systematically from those included, particularly if testing was unavailable because of severe disease or early mortality. The modest sample size in this cohort may also have limited statistical power to detect certain associations. The generalisability of the identified prognostic factors across other non-IPF ILD subtypes should be interpreted with caution, and potential class effects may not be fully represented. Finally, this single-centre tertiary referral cohort may not represent the wider Malaysian non-IPF ILD population. Larger prospective cohorts with standardised assessments and independent validation are needed to confirm these findings.

## Conclusion

In this multi-ethnic Malaysian cohort of non-IPF ILD, Indian patients were dominated. Older age and impaired baseline lung function were associated with higher mortality. The observed ethnic differences suggest that ethnicity warrants further investigation.

## Data Availability

All relevant data are within the manuscript and its Supporting Information files.

## Declarations

## Acknowledgements

I would like to thank my colleagues in the respiratory unit and the medical department of Universiti Malaya Medical Centre for their support to this study.

## Authors’ contributions

LCS: Study conceptualisation, data curation, formal analysis, writing the original draft, and manuscript review and editing. CKW: Supervision, data curation, writing the original draft, and manuscript review and editing. CKW, DS and DMC: Data curation. KYY, SYL, FBW and GYL: Data collection. WLL: Manuscript review and editing. MEP: Manuscript review. YKP: Supervision and study conceptualisation.

## Declaration on the use of AI

ChatGPT (OpenAI) was used for language editing, readability, grammar and assistance with reference checking.

## Data availability

The data are not publicly available because of patient confidentiality and institutional restrictions. Requests for access to de-identified data may be directed to the corresponding author and are subject to institutional and ethics approval.

## Supporting Information

**S1 Table.** Univariate analysis with FVC < 60% as the threshold.

| Variable | P value | Hazard ratio | 95.0% CI for Exp(B) |  |
| --- | --- | --- | --- | --- |
|  |  |  | Lower | Upper |
| FVC <60% of predicted | 0.134 | 0.570 | 0.273 | 1.189 |

**S2 Table.**
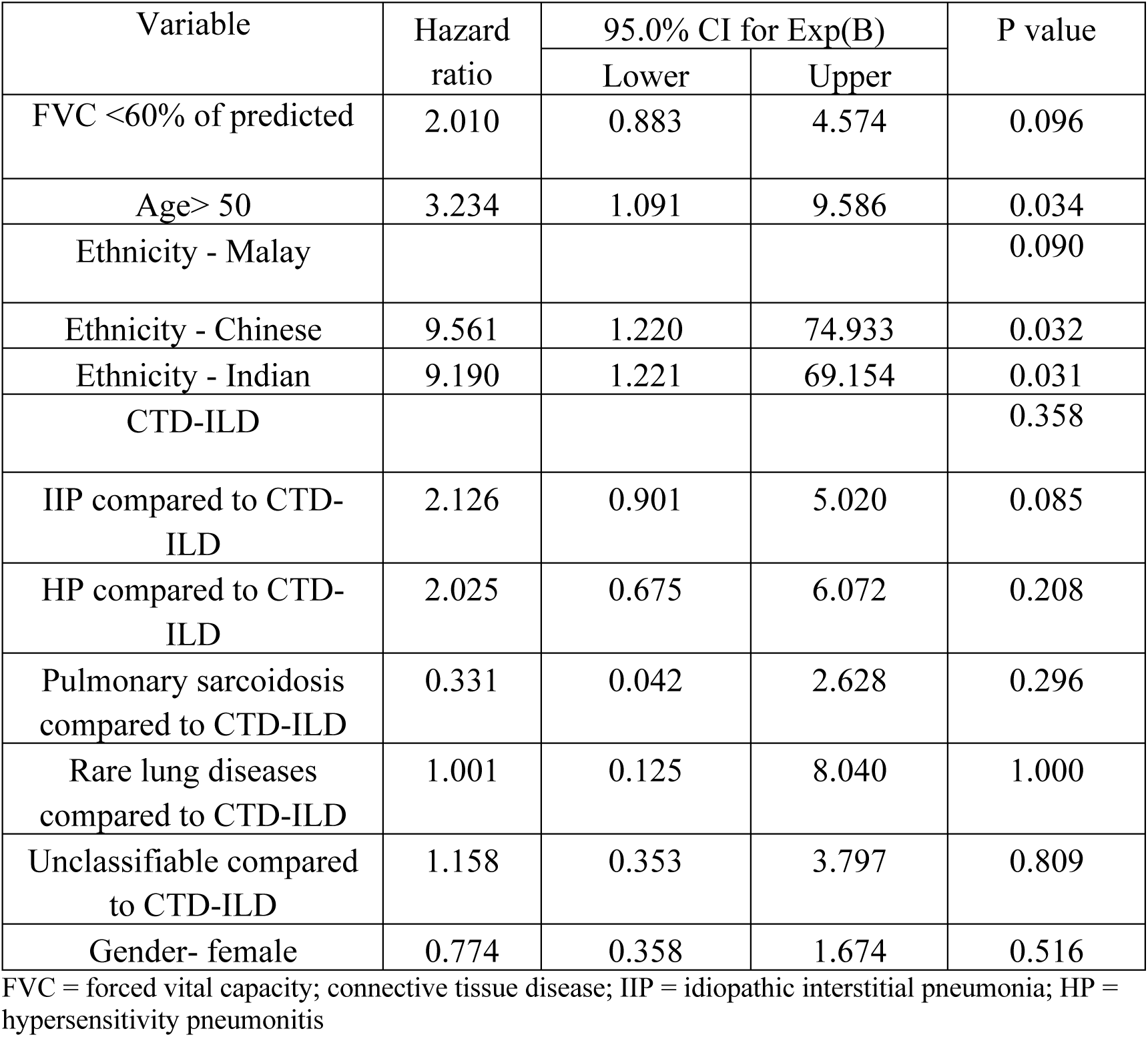
Multivariate analysis with FVC < 60% as the threshold.

| Variable | Hazard ratio | 95.0% CI for Exp(B) |  | P value |
| --- | --- | --- | --- | --- |
|  |  | Lower | Upper |  |
| FVC <60% of predicted | 2.010 | 0.883 | 4.574 | 0.096 |
| Age> 50 | 3.234 | 1.091 | 9.586 | 0.034 |
| Ethnicity - Malay |  |  |  | 0.090 |
| Ethnicity - Chinese | 9.561 | 1.220 | 74.933 | 0.032 |
| Ethnicity - Indian | 9.190 | 1.221 | 69.154 | 0.031 |
| CTD-ILD |  |  |  | 0.358 |
| IIP compared to CTD-ILD | 2.126 | 0.901 | 5.020 | 0.085 |
| HP compared to CTD-ILD | 2.025 | 0.675 | 6.072 | 0.208 |
| Pulmonary sarcoidosis compared to CTD-ILD | 0.331 | 0.042 | 2.628 | 0.296 |
| Rare lung diseases compared to CTD-ILD | 1.001 | 0.125 | 8.040 | 1.000 |
| Unclassifiable compared to CTD-ILD | 1.158 | 0.353 | 3.797 | 0.809 |
| Gender- female | 0.774 | 0.358 | 1.674 | 0.516 |

FVC = forced vital capacity; connective tissue disease; IIP = idiopathic interstitial pneumonia; HP = hypersensitivity pneumonitis

